# Phosphatidic Acid Mediated Sonodynamic Therapy Facilitates M1 Macrophage Osteoclastic Function and Alleviates Early Vascular Calcification

**DOI:** 10.1101/2024.10.10.24315275

**Authors:** Penghao Gao, Chao Zhao, Zixin Zhang, Qipin Zhou, Zhiyi Yang, Fanshuo Yin, Jialong Li, Yongxing Jiang, Xuezhu Zhao, Jiemei Yang, Tianyi Zhang, Linxin Wang, Qian Luo, Ye Tian

## Abstract

**BACKGROUND:** Vascular calcification significantly influences the onset and outcome of cardiovascular events, yet no effective treatment currently exists. Dysfunction of osteoclastic macrophages contributes to the formation of calcification. Our previous studies have shown that sonodynamic therapy (SDT) can rapidly reverse atherosclerotic plaques by targeting macrophages. This study aimed to investigate the effect of SDT on reducing early or mild vascular calcification by modulating the function of osteoclastic macrophages.

**METHODS:** Thirty-two patients with symptomatic femoropopliteal peripheral artery disease (PAD) were recruited to evaluate changes in vessel CT values and the target-to-background ratio (TBR) using positron emission tomography/computed tomography (PET/CT) 30 days post-SDT. An early calcification model was established in ApoE-/- mice, followed by SDT intervention. Frozen plaque sections from the mice were collected for mass spectrometry imaging (MSI)-based spatial metabolic analysis in situ. The NHGRI-EBI GWAS Catalog database and the human single-cell eQTL database (scQTLbase) were employed to analyze the causal relationship between key enzyme genes involved in phosphatidic acid (PA) synthesis in macrophages and vascular calcification using two-sample Mendelian randomization. To investigate cell ossification, calcification, and underlying mechanisms, RAW264.7 mouse macrophages were treated with a medium containing receptor activator of nuclear factor kappa-B ligand (RANKL), while mouse aortic vascular smooth muscle cells (MOVAS cells) were exposed to a calcification medium.

**RESULTS:** SDT significantly reduced the number of mildly calcified sites and the target-to-background ratio (TBR) of these sites in patients with femoropopliteal peripheral artery disease (PAD). In ApoE-/- mice, SDT alleviated early calcification of atherosclerotic plaques. MSI revealed that SDT altered the composition and distribution of lipid metabolites in atherosclerotic plaques, notably increasing the content of PA in the early calcified regions. Analysis of single-cell sequencing databases showed that key enzyme genes involved in PA synthesis—PLD1, PLD3, AGPAT4, and diacylglycerol kinase E (DGKE)—were enriched in macrophages of human coronary artery plaques. Mendelian randomization analysis indicated that DGKE negatively regulated coronary artery calcification. In vitro studies demonstrated that PA mediates SDT to promote M1 macrophage fusion and enhance carbonic anhydrase II (CA2) expression, thereby improving osteoclastic function and alleviating early calcification of MOVAS cells via the reactive oxygen species (ROS)-DGKE-PA pathway. In vivo, the CA2 inhibitor acetazolamide impaired the effects of SDT and exacerbated early calcification of atherosclerotic plaques in ApoE-/- mice.

**CONCLUSION:** This study demonstrates that PA-mediated SDT promotes M1 macrophage fusion and CA2 expression, improving osteoclastic function and alleviating early calcification through the ROS-DGKE-PA pathway.

**REGISTRATION:** URL: https://www.clinicaltrials.gov; Unique identifier: NCT03457662.

**What Is New?:**

1. SDT reduces early calcification in patients with symptomatic femoropopliteal PAD and in ApoE-/- mouse models of early calcification.
2. SDT upregulates the expression of PA in the early calcified regions of ApoE-/- mouse models.
3. PA-mediated SDT facilitates M1 macrophage fusion and enhances CA2 expression, thereby improving osteoclastic function and alleviating early calcification through the ROS-DGKE-PA pathway.

**What Are the Clinical Implications?:**

1. Targeting M1 macrophage DGKE-PA may serve as a potential intervention for treating early vascular calcification.
2. The combination of MSI and Mendelian randomization analysis proves to be an effective method for exploring key signaling lipids in disease.
3. PA-mediated SDT represents a promising approach for the effective reduction of early vascular calcification.

## INTRODUCTION

Vascular calcification refers to the ectopic deposition of hydroxyapatite in blood vessels^1,2^. The calcification within atherosclerotic plaques is involved in both plaque stabilization and rupture^3^. Early atherosclerotic plaques with microcalcification, as observed in pathology, carry a higher risk of rupture^4,5^. Mild calcification (low calcium density) detected via CT scans is associated with an increased risk of cardiovascular disease^6–8^. Typically, early atherosclerotic plaques exhibit mild calcification on CT^8^. Furthermore, intensive lipid-lowering therapy with statins has not been shown to prevent the progression of calcified aortic stenosis or induce its resolution^9^. In fact, statins have been found to promote the progression of calcification in coronary atherosclerosis^10^. Anti-inflammatory therapy using IL-1β antibodies has proven ineffective against the calcification of advanced atherosclerotic plaques in Apoe-/- mice^11^. To date, no effective therapy exists for early or mild calcification.

For decades, vascular calcification was regarded as a passive and degenerative process. However, accumulating evidence suggests that vascular calcification, akin to bone mineralization, is an actively regulated process involving both induction and inhibition mechanisms^12,13^. Multinucleated osteoclast-like cells, derived from macrophages, are present in human atherosclerotic plaques^14^. These osteoclastic macrophages surrounding calcification play a crucial role in mitigating calcification^15^. Nevertheless, these cells are not fully differentiated and exhibit dysfunction^16^. Currently, therapies targeting the regulation of osteoclastic macrophages in vascular calcification face significant challenges and have yet to be tested in animals or humans^15^.

Lipids play a crucial role in the induction and inhibition of atherosclerotic calcification. During the calcification process, extracellular vesicles, including exosomes and apoptotic bodies, are released. These local extracellular vesicles are rich in cholesterol, diacylglycerol, sphingomyelin, ceramides, and other substances, providing microcalcification nucleation sites composed of phosphatidylserine and membranin^17^. Calcium and inorganic phosphorus (Pi) can accumulate within these extracellular vesicles, where they develop into amorphous calcium phosphate and ultimately form microcalcified crystal structures such as hydroxyapatite^17,18^. Phosphatidic acid, one of the simplest membrane phospholipids, is an important signaling molecule involved in regulating various cell signaling pathways and functions, as well as membrane rearrangement18. Previous studies have shown that PA is regulated by ROS^19,20^, and that PA can promote macrophage fusion and enhance osteoclastic function^21^. However, there are currently no methods for the targeted regulation of PA in macrophages in vivo, and no studies have investigated the effect of PA on vascular calcification through osteoclast regulation.

SDT primarily activates sonosensitizers to produce brief and moderate ROS, playing multiple roles in the process^22,23^. We have conducted a series of studies on SDT targeting macrophages in atherosclerotic plaques. Experimental studies demonstrated that SDT could promote cholesterol efflux, elicit an anti-inflammatory response, facilitate the transformation of M1 macrophages into M2 macrophages, and induce autophagy in macrophages^24,25^. Translational studies confirmed that SDT significantly reduced arterial inflammation, angiogenesis, and the rate of vascular stenosis in carotid and femoral artery plaques in both animal models and human patients after one month^26,27^. A randomized controlled clinical trial indicated that SDT could lower inflammation levels and improve functional performance in patients with lower extremity arterial occlusions^28^.

In the present study, we investigated the effects of SDT on early vascular calcification and found that SDT significantly reduced early vascular calcification in both patients and mice. MSI revealed that SDT upregulated the expression of PA in the early calcified regions. Mendelian randomization analysis of single-cell sequencing indicated that DGKE negatively regulates coronary artery calcification. Furthermore, animal and cellular experiments demonstrated that PA mediates SDT-induced fusion of M1 macrophages and enhances CA2 expression, improving osteoclastic function and alleviating early calcification through the ROS-DGKE-PA pathway

## METHODS

### Animal experiment

The animal protocol was approved by the Ethics Committee of Harbin Medical University. All experiments were performed in accordance with the ethical guidelines set out by the United Kingdom Animals (Scientific Procedures) Act 1986 and Directive 2010/63/EU of the European Parliament on the protection of animals used for scientific purposes. Studies conformed to the Guide for the Care and Use of Laboratory Animals published by the U.S. National Institutes of Health. Male ApoE-/- mouse C57BL6 background (Qingjilan Technology Co., LTD., Nanjing, China) were raised at the Animal Care Center of the First Affiliated Hospital of Harbin Medical University. The flow diagram of animal experiments is shown in Figure S1, and Expanded methods are available in the Supplemental Appendix.

### Cell experiment

Mouse RAW264.7 macrophages (Otwo Biotechnology Corporation, Shenzhen, China), and MOVAS cells (Otwo Biotechnology Corporation, Shenzhen, China) were cultured in a humidified atmosphere with 5% CO2 at 37℃. Detailed cell experiments methods are available in the Supplemental Appendix.

### Clinical trial

The SMART-PAD is an investigator-initiated, phase-2, single center, prospective, randomized, double-blind, sham-controlled, and parallel-designed clinical trial. This trial was performed at the First Affiliated Hospital of Harbin Medical University, China. Participants were assigned to either the SDT group or the sham control group. The trial was approved by the Institutional Review Board of Harbin Medical University, in compliance with the principle of the Declaration of Helsinki. All participants gave their written informed consent. The CONSORT flow-diagram is shown in Figure S2, and the detailed clinical trial methods are available in the Supplemental Appendix.

### SDT intervention

Sonosensitizer for SDT in this study was Sinoporphyrin sodium (DVDMS). DVDMS is the property of Qinglong Hi-tech Co.,Ltd.(Jiangxi,China) and was provided by Professor Qicheng Fang from the Chinese Academy of Medical Sciences(Beijing,China). The ultrasound device was manufactured by Harbin Institute of Technology (Harbin, China). The ultrasonic transducer parameters were as follows: diameter is 35 mm, and resonance frequency is 1.0 MHz. DVDMS was used as the sonosensitizer for SDT. The animals were kept away from light during and after SDT treatment for 24 h. At 4 hours after intravenous injection of DVDMS (4mg /kg) administration, anesthetized animals were subjected to ultrasound for 15 min with an ultrasonic intensity of 0.4 W/cm^2^ for mice, as previously described ^27, 29^. The ultrasound was applied as previously described^25^. In brief, for mice, the ultrasonic transducer was placed under the chest through a degassed water column. For the in vitro study, cells were plated on 35-mm Petri dishes that were placed on a degassed water bath 30 cm away from the ultrasonic transducer. Based on the optimized SDT parameters, RAW264.7 macrophages were incubated with 0.8μM DVDMS for 4 h, followed by irradiation with an ultrasound intensity of 0.2 W/cm^2^ for 7 min.

According to randomization results, 0.05 mg/ml of DVDMS solution or normal saline as placebo was administrated intravenously at a dosage of 0.2 mg/kg body weight to the participants. Therapeutic ultrasound exposure (35-mm 1 MHz transducer, Harbin Institute of Technology, Harbin, China) was performed after 4 hours after DVDMS or placebo was administered. The corresponding skin points of the targeted lesions were exposed to therapeutic or pseudo ultrasound for 15 min per point with 2.1 W/cm^2^ of ultrasound intensity to plaques in the common femoral artery (CFA) and superficial femoral artery (SFA), and 1.8 W/cm^2^ to plaques in the popliteal artery (POA) of both legs.

### Mass spectrometry imaging

ApoE-/- mice at 6 weeks of age were fed an atherosclerotic diet (22% fat, 0.12% cholesterol), and ApoE-/- mice in the control group and SDT group were fed a high-fat diet for 22 weeks with atherosclerotic plaques collected at the aortic root. The plaque samples from different three mice were immediately washed with ice brine and frozen in liquid nitrogen for 1 min at −80℃. Frozen sections (14 μm/ sheet) were cut in a freezer and transferred to slides coated with ITO and stored at −80℃ for MSI and histochemical staining. Before the experiment, dry it in a dryer for 40 minutes.

The matrix solution for MALDI-MSI, 2.5-dihydroxybenzoic acid (≥99.0% HPLC, Sigma-Aldrich, USA), was prepared at a concentration of 40.0 mg/mL in H_2_O (v:v=1:1). It was then applied using the TM Sprayer (HTX Technologies, USA) with a nozzle temperature set to 60 °C, pressure maintained at 10 psi, solvent flow rate of 0.1 mL/min, and nozzle velocity adjusted to 800 mm/min. MALDI-TOF/TOF MSI (Bruker UltrafleXtreme, Bruker, Germany) was used for MSI data acquisition. The detailed instrumental parameters of MSI included the m/z 500-900 in positive ionization modes, 50% of laser power, 20 kV of ion source voltage, 20 kV of reflector voltage and 50 μm of spatial resolution.

The image data acquired from MALDI-MSI were imported into SCiLS software and FlexImaging software (Bruker, Germany) to perform image segmentation, multivariable statistics (probabilistic latent semantic analysis with random initialization for component analysis), lipid screening and visualization, and quantitative analysis. Meanwhile, regions and sub-regions of samples were annotated by using histomorphologic spectra, including vascular lumen area, myocardium area, extravascular tissue area, early atherosclerotic calcification area, atherosclerotic area, and valvular area.

### Single cell sequencing and Mendelian randomization

The scRNA-seq dataset for patients with acute coronary syndromes (ACS) was downloaded to the GEO database with entry number GSE184073, which was generated by the 10x platform. The dataset consisted of 1,720 cells from three patients. After excluding the low-quality cells whose nFeature_RNA was significantly lower than other clusters, we performed principal component analysis on the scaled data, and used the first 30 principal components to perform for graph-based clustering to identify cell clusters. Ultimately, our analysis yielded a total of 1,531 cells in all patients included in this study.

Mendelian randomization analysis was performed using the “TwoSampleMR” software package in R. Inverse variance-weighted (IVW) meta-analysis was chosen as the primary Mendelian randomization analysis method because of its best accuracy and good IV quality. Various auxiliary methods such as MR Egger, weighted median, simple model and weighted model were used to evaluate the causal relationship. The Mendelian randomization analysis flow diagram is shown in Figure S3.

Sensitivity analysis. To further ensure the quality of the instrumental variable (IV), an extensive sensitivity analysis was performed. Heterogeneity among IVs was assessed by Cochran’s Q test. MR Pleiotropic residual and outlier (MR-PRESSO) method was used to detect MR Pleiotropic, in which the global test detects horizontal pleiotropic [29686387]. In addition, a leave-one-out analysis was performed to identify potentially impactful SNPS that had a significant impact on the remaining IVW results.

### Statistical Analysis

All experiments were repeated three or more times, and the data were analyzed by statisticians who were double-blind to the experiments. Statistical analysis was performed using Graphpad Prism 9.0 (La Jolla, USA) software. Counting data are expressed as frequency and percentage, and Fisher’s exact test or x2 test is used between groups. Measurement data are expressed as mean ± standard deviation. Comparison between groups Shapiro-Wilk tested the normality of the data. If the data met the normal distribution, the paired T-test was used for comparison within the group and the independent sample T-test was used for comparison between two groups. For non-normal distribution, wilcoxon test was used for paired samples and Mann-Whitney test was used for two independent samples. If the correlation analysis meets the normal distribution, Pearson correlation test is used. If the normal distribution is not satisfied, Spearman test is used. One-way analysis of variance (ANOVA) was used for comparison among multiple groups, and Kruskal-Wallis test was used for comparison among multiple groups if the data did not meet the normal distribution. P<0.05 was statistically significant.

## RESULTS

### SDT reduces the number of mildly calcified sites and their inflammation in PAD patients

To explore the effect of SDT on mildly calcified sites, a total of 139 patients were screened at the First Affiliated Hospital of Harbin Medical University between March 15, 2018, and November 13, 2018. Of these patients, 32 were enrolled and randomly assigned to the control or SDT groups. The most common reason for exclusion was the ABI higher than 0.9 (33% of the excluded patients). The primary and secondary endpoints of this clinical trial have been published in the paper which focus on arterial inflammation^28^. Basic patient information, laboratory tests, and adverse events are also provided in the published paper^28^. At present, we will mainly present the exploratory endpoints of this clinical trial which focus on vascular calcification.

At baseline, the mean age of participants was 64.7 years, 63% were male, 56% were current smokers, and 59% had diabetes. The SDT and control groups were well matched. A total of 113 targeted lesions in the index-leg were treated, of which 60 were treated with SDT and 53 with pseudo SDT. The median number of lesions per patient was 3.5, ranging from one to five. The characteristics of the lesions were matched between groups.

Exploratory Endpoints: All participants in the SDT group and 15 (94%) participants in the control group completed the PET/ CT scan at the 30-day follow-up. Four participants in the SDT group and three participants in control group were excluded from the analysis due to the lack of calcified plaque. Representative cross-sectional CT, 18F-FDG-PET and fused 18F-FDG-PET/CT images before and after SDT (Figure 1A-B). PET/CT results showed that compared with the control group, SDT significantly reduced the number and the percentage of mildly calcified sites from 375 (15.8%) to 331 (13.9%) (Figure 1C-E). While, SDT did not reduce the number and the percentage of severely calcified sites from 366 (15.4%) to 342 (14.4%) (Figure 1I). Compared with the control group, SDT significantly reduced the TBR values of mildly calcified sites(Figure 1G). But, SDT did not reduce the TBR value of the severely calcified sites (Figure 1K). Compared with the control group, SDT did not reduce the CT values of mildly and severely calcified sites (Figure 1F, J).

**Figure 1.**
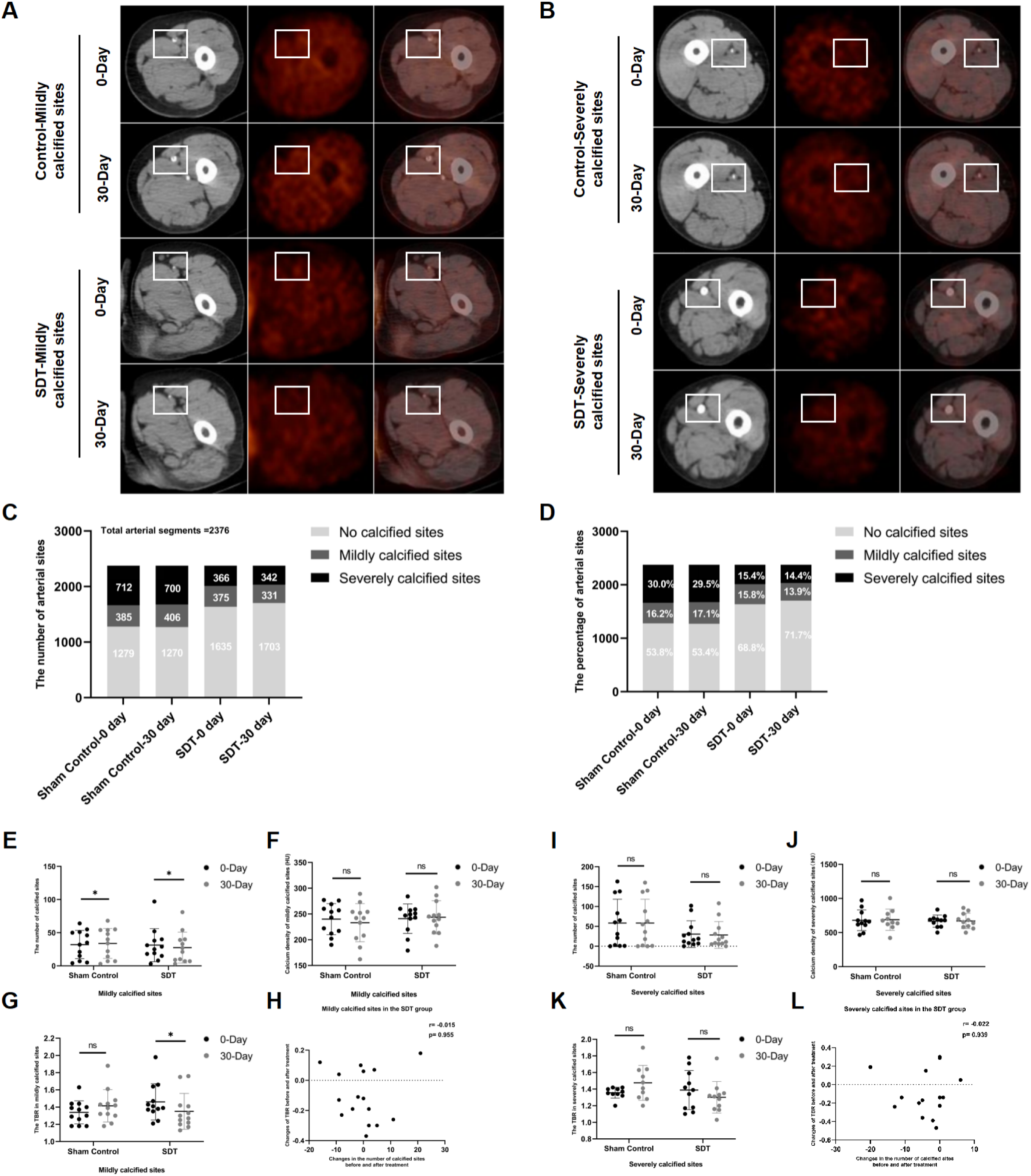
SDT reduces the number of mildly calcified sites and their inflammation in PAD patients. (A) Representative cross-sectional CT, 18F-FDG-PET and fused 18F-FDG-PET/CT images of the femoropopliteal artery with mild calcification at the same site in the same patient before and after SDT. The white box indicates the diseased femoropopliteal artery. (B) Representative cross-sectional CT, 18F-FDG-PET and fused 18F-FDG-PET/CT images of the femoropopliteal artery with severe calcification at the same site in the same patient before and after SDT. The white box indicates the diseased femoropopliteal artery. (C)The number of non-calcified sites (calcium density, < 130 HU), mildly calcified sites (calcium density, 130-399 HU), and severely calcified sites (calcium density,≥400 HU) changes respectively at baseline and 30 days after treatment in the control group and the SDT group. The number of total arterial sites n=2376. (D) The percentage of non-calcified sites (calcium density, < 130 HU), mildly calcified sites (calcium density, 130-399 HU), and severely calcified sites (calcium density, ≥400 HU) of total arterial sites changes respectively at baseline and 30 days after treatment in the control group and the SDT group. The number of total arterial sites n=2376. (E) The number of mildly calcified sites at baseline and 30 days after treatment in the control and SDT groups. Control group n = 12, SDT group n = 12. (F) Mean CT values of mildly calcified sites at baseline and 30 days after treatment in the control and SDT groups. Control group n = 12, SDT group n = 12. (G) TBR values of mildly calcified sites at baseline and 30 days after treatment in the control and SDT groups. Control group n = 12, SDT group n = 12. (H) Correlation between changes in the number of mildly calcified sites and changes in TBR value after SDT. Control group n = 12, SDT group n = 12. (I) The number of severely calcified sites at baseline and 30 days after treatment in the control and SDT groups. Control group n = 12, SDT group n = 12. (J) Mean CT values of severely calcified sites at baseline and 30 days after treatment in the control and SDT groups. Control group n = 12, SDT group n = 12. (K) TBR values of severely calcified sites at baseline and 30 days after treatment in the control and SDT groups. Control group n = 12, SDT group n = 12. (L) Correlation between changes in the number of severely calcified sites and changes in TBR value after SDT. Control group n = 12, SDT group n = 12.

Postmortem analysis: For mildly calcified sites, there was no correlation between the changes in the number of calcified sites and the changes in TBR values of calcified sites before and after SDT (Figure 1H). For severely calcified artery segments, there was no correlation between the changes in the number of calcified sites and the changes in TBR values of calcified sites before and after SDT (Figure 1L).

### SDT alleviates early aortic calcification in ApoE-/- mice

We further verify the therapeutic effect of SDT on calcification in an experimental ApoE-/- aortic atherosclerosis early calcification model in high-fat fed mice for 20 weeks. Compared with the control group, the ratio of bone morphogenetic protein 2 (BMP2), osteocalcin (OST) immunofluorescence positive area to plaque area was significantly reduced in SDT group at the 15th day after SDT (Figure 3A-B, Figure S4B-C). But, there was no significant change in the percentage of BMP2, OST immunofluorescence positive area in the group 30 days after SDT compared to the control group (Figure 3A-B, Figure S4B-C). The percentage of BMP2 immunohistochemically positive area was significantly lower in the group 15 days after SDT compared to the control group. But, there was no significant change in the percentage of BMP2 immunohistochemically positive area in the group 30 days after SDT compared to the control group. This suggests that SDT can significantly reduce calcified area at the 15th day after SDT. (Figure 2C-D). The relative calcified volume of the proximal or distal aortic segment in micro-CT was significantly reduced at the 15th day after SDT compared to the control group (Figure 3E-F).

**Figure 2.**
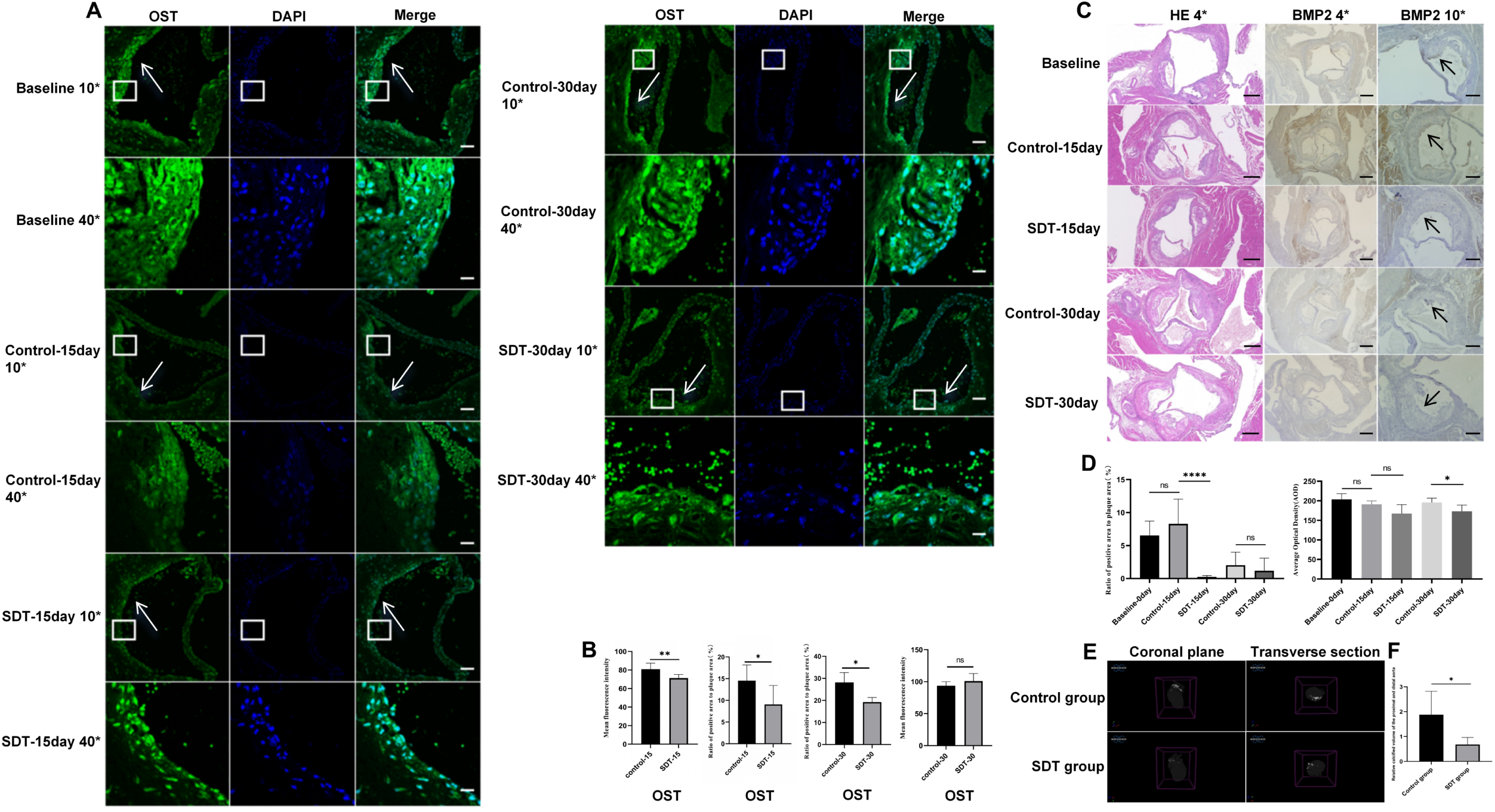
SDT alleviates early aortic calcification in ApoE-/- mice. (A) Representative OST immunofluorescence staining of aortic root atherosclerosis in ApoE-/- mice before and after SDT (10* scale =200um, 40* scale =50um). (B) The percentage of OST positive area to the total plaque area and the mean fluorescence intensity, n = 7 in the Control-15day group, n = 6 in the SDT-15day group, n = 4 in the Control-30day group, n = 4 in the SDT-30day group. (C) Representative HE and BMP2 histochemical staining of atherosclerotic calcification in ApoE-/- mice aortic root before and after SDT, scale =500um. (D) The percentage of BMP2 positive area in the total plaque area and the average optical density value. n = 10 in Baseline group, n = 4 in Control-15day group, n = 10 in SDT-15day group, and n = 6 in Control-30day group, n = 10 in SDT-30day group. (E) Micro-CT images of control group and SDT group. (F) The relative calcified volume of the proximal or distal aortic segment of the control group and the SDT group at the 15th day after SDT. n = 4 in control group and n = 4 in SDT group.

**Figure 3.**
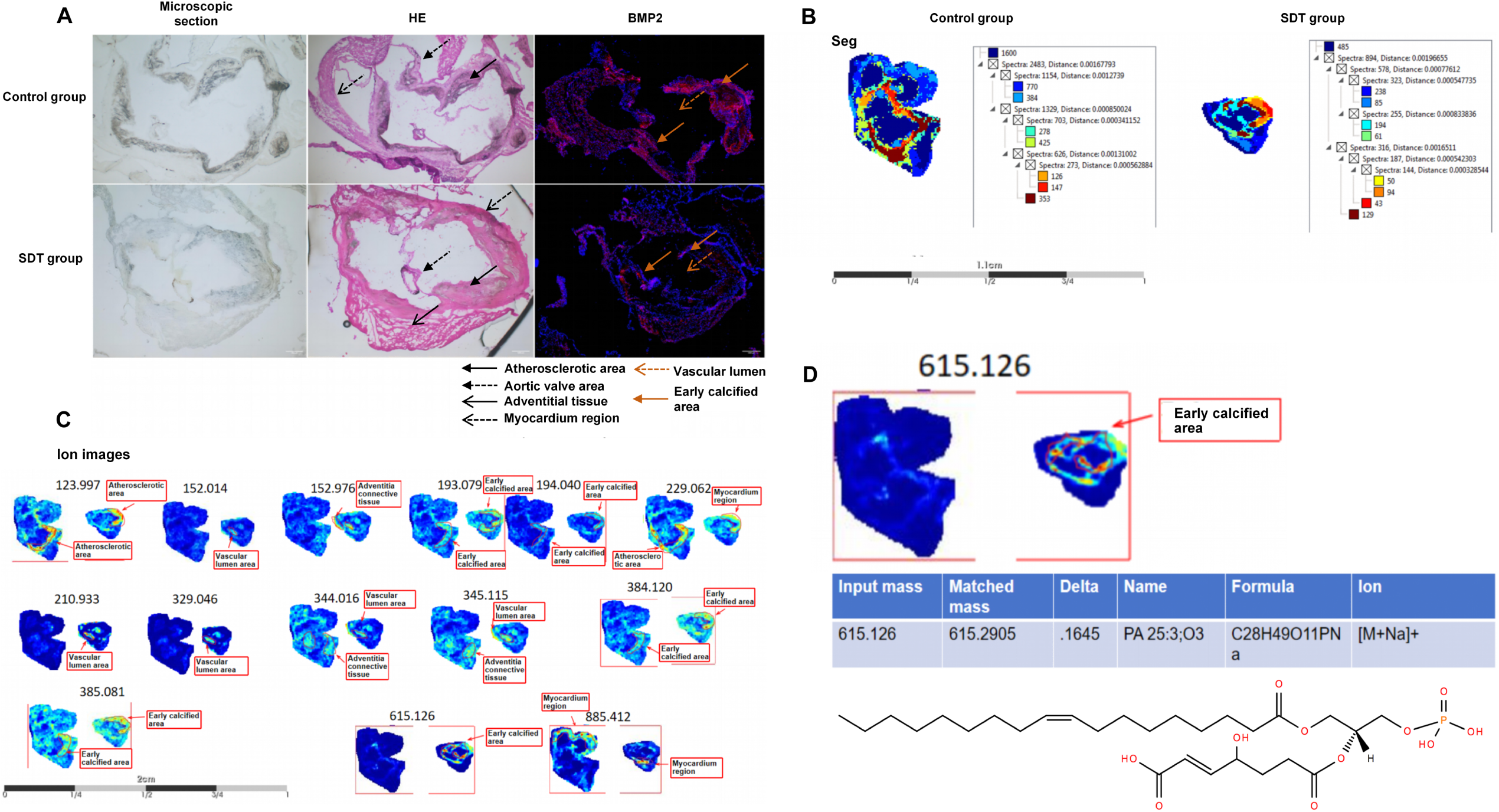
SDT increases the expression of PA in early calcified areas of ApoE-/- mice. (A) HE staining and BMP2 immunofluorescence staining images of serial frozen sections of aortic root plaques of mice in SDT group and control group and the division of different areas in plaques. (B-C) MALDI-MSI images of spatial distribution of plaques in aortic root of mice in SDT group and control group. (D) PA(25:3) m/z:615.126 distributed in the early calcified area of the aortic root of mice and is significantly up-regulated in the SDT group.

### SDT increases the expression of PA in early calcified areas of ApoE-/- mice

Lipid metabolism is not only deeply involved in the calcification process^17,18^, but also regulates the polarization and function of macrophages^30,31^. We applied mass spectrometry imaging (MSI) to explore the mechanisms underlying SDT. We obtained 12-14μm sections of early calcified plaques from the aortic root of one mouse in the SDT group and another in the control group. After MALDI scanning, adjacent sections were stained with HE reagent and BMP2 immunofluorescence (Figure 3A). We also described the precise regions before and after SDT. The lipid spatial profile of early calcified plaques changed before and after SDT, which could be divided into vascular lumen area, atherosclerotic area, valvular area, connective tissue area, myocardial area, and early calcification area (Figure 3B-C). Focusing on the early calcification area, we found that PA(25:3) m/z:615.126 was distributed in the early calcification area of the mouse aortic root and was significantly up-regulated in the SDT group at the 15th day after SDT. (Figure 3D).

### The DGKE of monocyte/macrophages negatively regulates coronary atherosclerotic calcification

PA has a variety of pathophysiological functions, including involvement in the regulation and expansion of numerous cell signaling pathways, membrane rearrangement, and the regulation of cell proliferation, survival, cytoskeletal organization, vesicle transport, and secretion^19^. However, the redundancy and multilevel regulation of PA metabolism, along with the ability of PA biosynthetases to partially compensate for one another, complicate the study of the biological functions of specific PA pools^32^.

To further investigate whether SDT affects PA metabolism and the effect of PA pools on calcification, we analyzed single cells from 3 patients with ACS using a 10x scRNA-seq platform. After the standard single-cell analysis process, a total of 1531 cells were obtained. We used uniform manifold approximation and projection (UMAP) methods to identify five major clusters, including T cells (n = 732; CD3D, CD3E, NKG7), plasma cells (n = 42; MZB1, IGHG3, JCHAIN), monocyte (n = 445; FCN1, S100A8, S100A9, S100A12), c1q_trem_ macrophages (n = 181; TREM2, C1QC, APOE), B cells (n = 131; CD79A CD19)(Figure 4A-B). Expressions of key PA synthesis enzyme genes PLD1,PLD3,AGPAT4 and DGKE were found in mononuclear macrophages of coronary plaque (Figure 4C).

**Figure 4.**
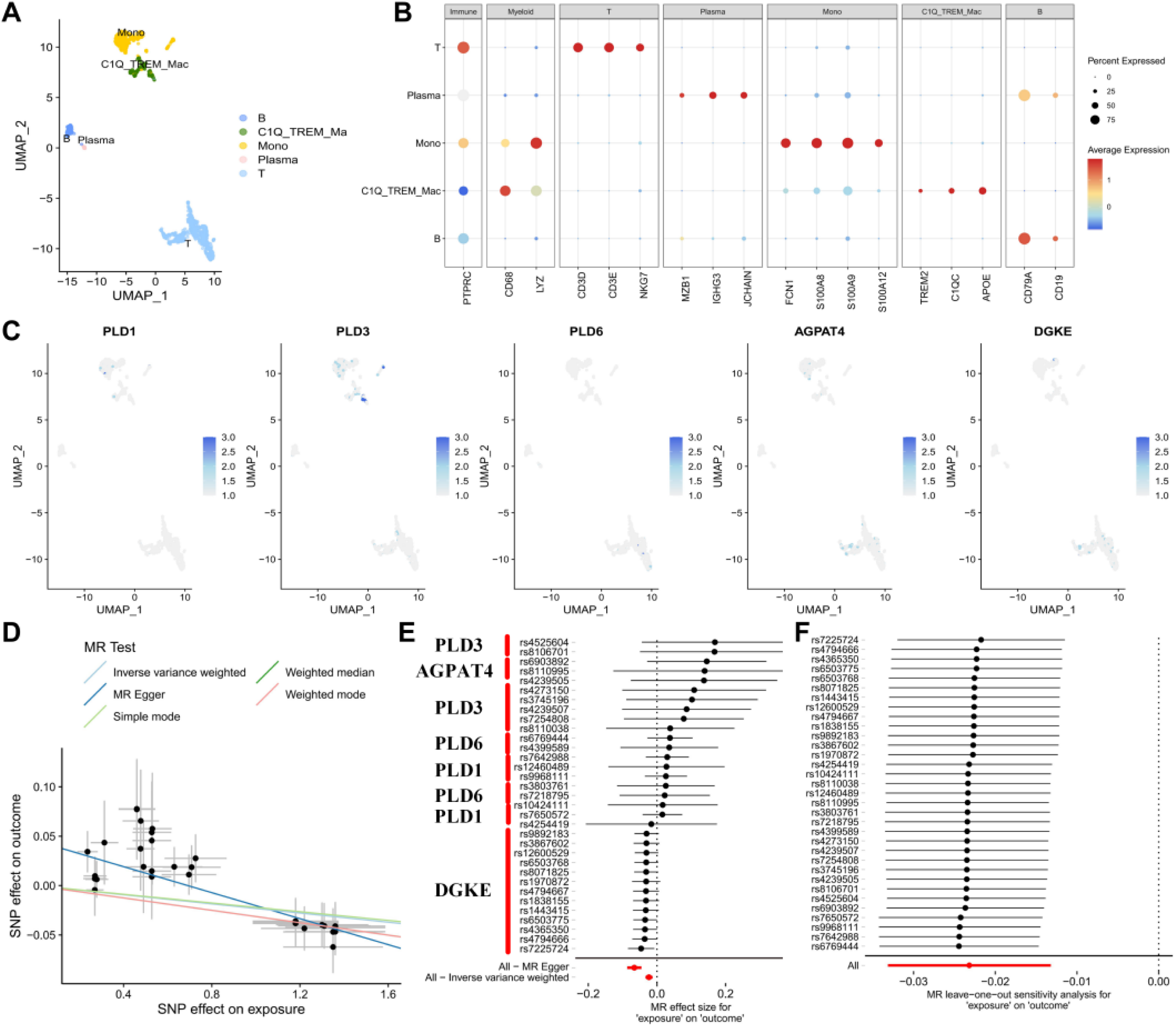
The DGKE of monocyte/macrophages negatively regulates coronary atherosclerotic calcification. (A) Uniform manifold approximation and projection of coronary plaque cells. (B) Gene expression of coronary plaque cell subsets. (C) Specific expression of key enzymes of PA synthesis in monocyte/macrophages of coronary artery plaques. (D) Scatter plot of the causal effect of SNPS of key enzyme genes in PA synthesis on coronary artery calcification disease. (E) Forest plot of associations between SNPS of key enzyme genes for PA synthesis and coronary artery calcification disease. (F) leave-one-out analysis of SNPS in key enzyme genes of PA synthesis.

We extracted the genes and eQTL for key enzymes of PA synthesis in monocyte/macrophage cells from scQTLbase^33^, and focused our study on significant SNPs (P < 1e^-6^) within 1Mb distance from gene starting point and endpoint as instrumental variables of exposure factors. A study from the Cohorts for Heart and Aging Research in Genomic Epidemiology Consortium (CHARGE), which included 26,909 individuals of European ancestry and 8,867 individuals of African ancestry, was obtained from the GWAS Catalog database and was the largest GWAS meta-analysis of multi-ancestral coronary artery calcification to date. We treated this as a calcification result and performed Mendelian randomization analysis (GCST90278455). It was found that IVW estimated the risk of vascular calcification decreased with the increase of PA synthase level (or = 0.9770839,95% confidence interval CI: 0.97-0.99, P = 5.091079 e^-6^). The five models (IVW, MR Egger, weighted median, simple model and weighted model) showed the same direction. This can be demonstrated by the negative slope of all lines of PA synthetase levels and coronary artery calcification results (Figure 4D). Forest plots show that high PA synthetase levels are protective factors for coronary artery calcification, and DGKE plays a major protective role (Figure 4E). Cochran’s Q test found no heterogeneity in our study. MR-Egger regression showed that there was a pleiotropy between PA synthetase level and coronary artery calcification results (P = 6.621183e^-5^). However, pleiotropy was not detected with the MR-PRESSO global test and SNP was not removed (P = 0.419)(Figure S5). The “leave-one-out” also determined that no estimates of all significant exposure outcomes had a significant impact on the overall MR Analysis results (Figure 4F). All SNPs passed the correlation strength threshold because the minimum f statistic was 22.396, indicating that weak instrument bias was unlikely in the MR Analysis and the MR results in this study were relatively robust.

### SDT targeting M1 macrophages alleviates the calcification of MOVAS cells

Macrophages play a pivotal role in the regulation of vascular calcification process, and the pro-inflammatory/anti-inflammatory phenotype of macrophages directly affects the stage and size of calcification, but the phenotype of macrophages around early calcification is still unknown^34,35^. In the early calcification model of ApoE-/- mice fed high fat for 20 weeks, immunofluorescence staining showed that calcification related protein BMP2 and maker CD86 of M1 macrophages and marker CD206 of M2 macrophages were roughly in the same region (Figure 5A, Figure S4A). To study the effects of macrophages with different phenotypes on calcification formation, mouse RAW264.7 macrophages with different phenotypes were induced and MOVAS cells calcification model was established (Figure S6A-D,F-G). Mouse RAW264.7 macrophages with different phenotypes were directly co-cultured with MOVAS cells respectively, and the expressions of calcification related proteins BMP2,osteogenic transcription factor (RUNX2), and osteopontin (OPN) in the co-culture system were measured individualy. Compared with M1 macrophages, M2 macrophages could reduce calcification related protein expression in co-culture system (Figure 5C).

**Figure 5.**
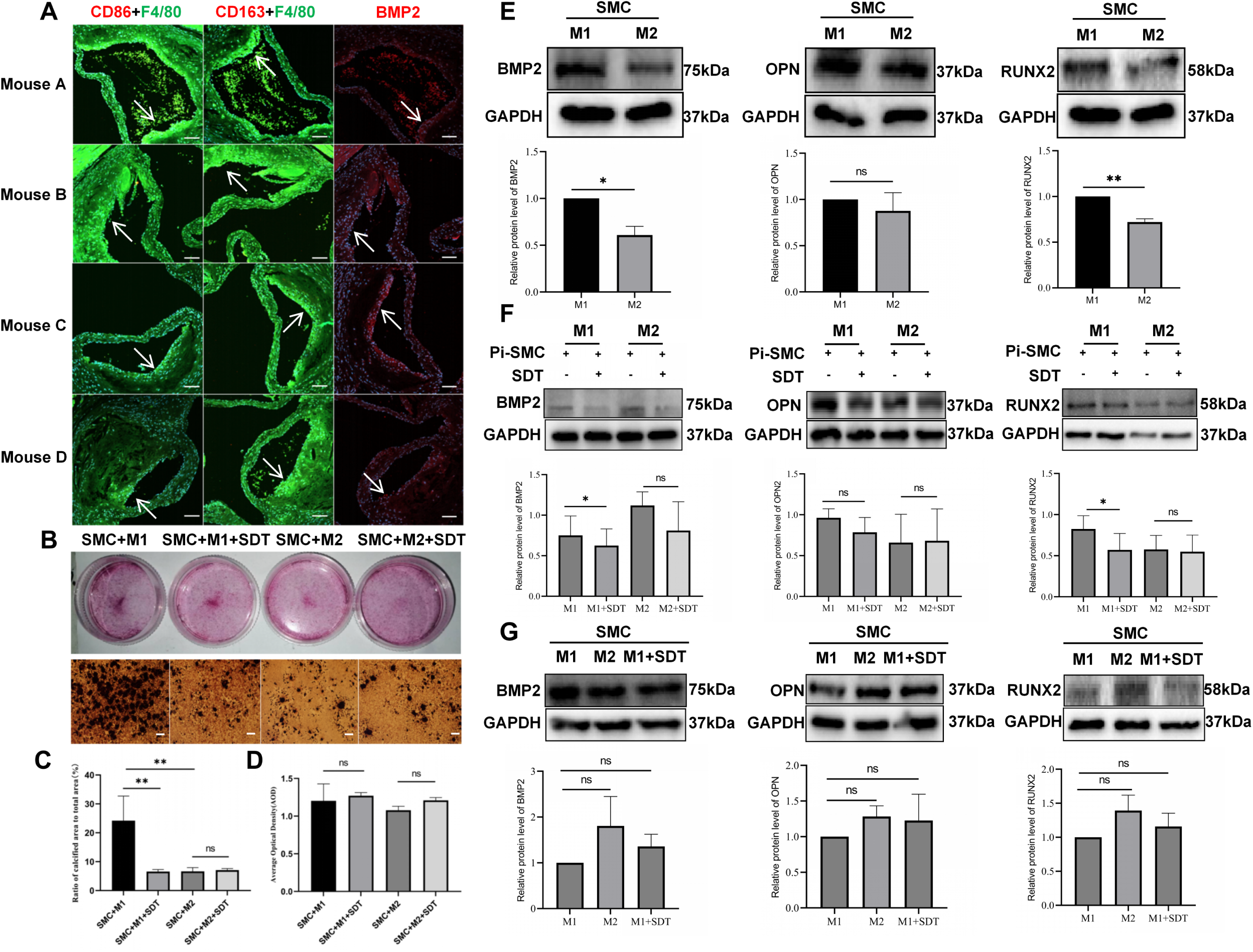
SDT targeting M1 macrophages alleviates the calcification of MOVAS cells. (A) Immunofluorescence staining representatives immunofluorescence staining of M1 macrophage marker CD86, M2 macrophage marker CD163 and calcification associated protein BMP2 (bone morphogenetic protein 2) in early calcification model of ApoE-/- mice. Scale =200um. (B) Alizarin red staining representatives of direct co-culture system of M1 macrophages or M2 macrophages and calcified MOVAS cells treated with or without SDT. Magnification of microscope field (×100). Scale =500um. n = 3. (C-D)Statistical graphs of direct co-culture system of M1 macrophages or M2 macrophages and calcified MOVAS cells treated with or without SDT. *P < 0.05, **P < 0.01. (E) Westernblot representatives and statistical graphs of calcification related proteins BMP2, OPN and RUNX2 in direct co-culture system of M1 macrophages or M2 macrophages and calcified MOVAS cells. n = 3. *P < 0.05, **P < 0.01. (F) Westernblot representatives and statistical graphs of calcification related proteins BMP2, OPN, RUNX2 in direct co-culture system of M1 macrophages or M2 macrophage and calcified MOVAS cells treated with or without SDT. n = 3. *P < 0.05. (G) Westernblot representatives and statistical graphs of calcification related proteins BMP2, OPN, RUNX2 in indirect transwell co-culture system of M1 macrophages or M2 macrophage and calcified MOVAS cells treated with or without SDT. n = 3. *P < 0.05.

To explore the effects of different phenotype macrophages treated with SDT on MOVAS cells calcification, M1 macrophages or M2 macrophages treated with SDT were directly co-cultured with calcified MOVAS cells respectively. The expressions of calcification related proteins BMP2, OPN, RUNX2 in M1 macrophages treated with SDT and calcified MOVAS cells directly co-culture system were lower than those in non-SDT relative co-culture system. SDT had no obvious effects on M2 macrophages relative co-culture system (Figure 5D). Alizarin red staining also showed that SDT could reduce the calcification of M1 macrophages and calcified MOVAS cells directly co-culture system, but SDT had no obvious effects on M2 macrophages relative to the co-culture system (Figure 5B). SDT had no obvious effects on the macrophage-MOVAS cells indirect Transwell co-cultured system (Figure 5E). Our results indicated that SDT reduced the calcification of M1 macrophage-MOVAS cells co-culture system depending on the intercellular contact.

### SDT reduces the calcification of MOVAS cells through ROS-DGKE-PA pathway

Flow cytometry showed that MOVAS cells did not phagocytose sonosensitizer, while macrophages significantly phagocytose sonosensitizer after incubating DVDMS for 4 hours in vitro identifing the targeting cells of SDT were macrophages (Figure 6A). RNA sequencing showed that SDT changed significantly the lipid metaboly-related pathways in M1 macrophages treated with SDT, indicating that SDT significantly altered lipid metabolism in M1 macrophages (Figure 6B).

**Figure 6.**
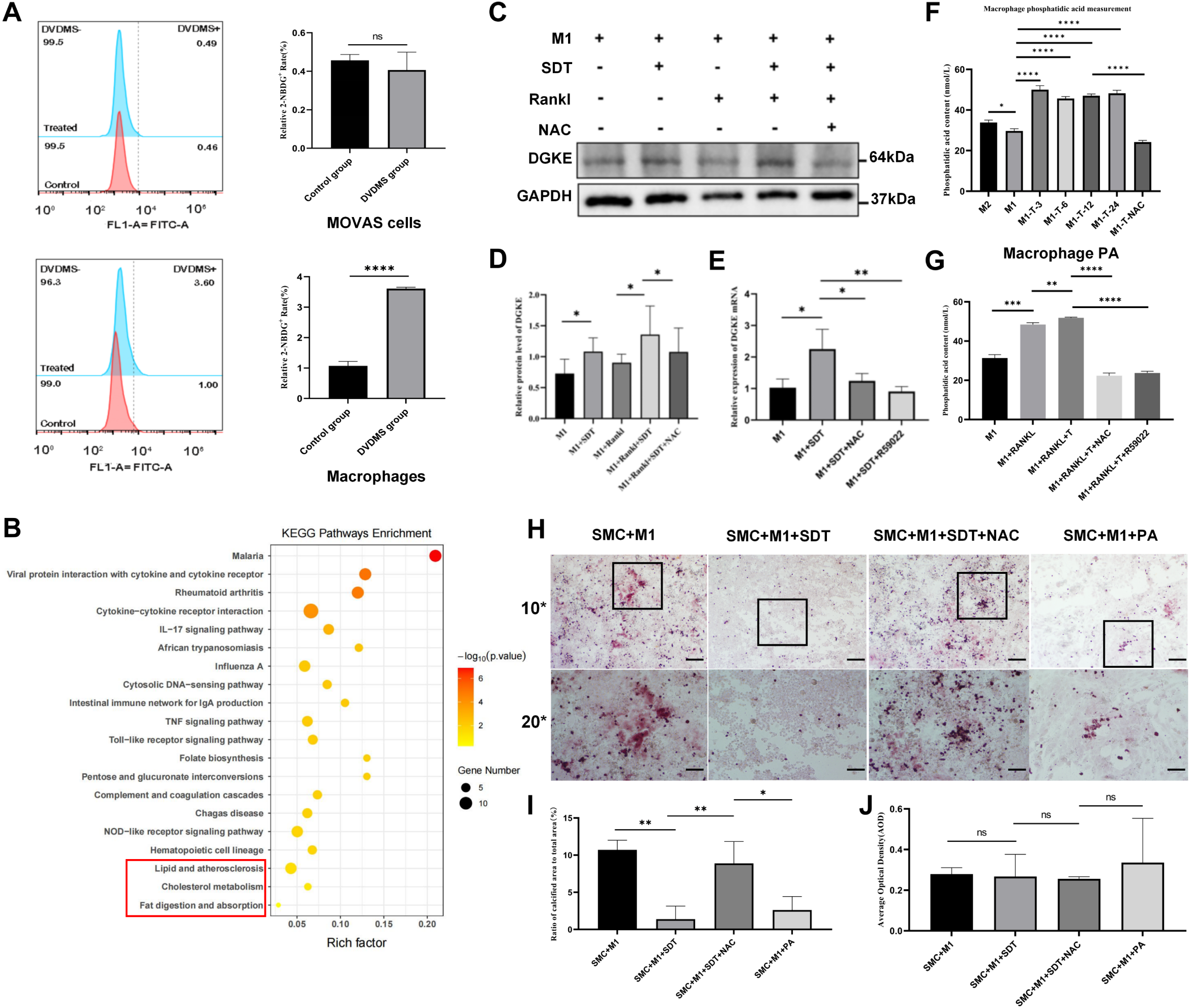
SDT reduces the calcification of MOVAS cells through ROS-DGKE-PA pathway. (A) Flow cytometry representatives and statistical graphs of MOVAS cells and macrophages treated with DVDMS for 4 hours, respectively. (B) The transcriptional expression profiles of M1 macrophages and M1 macrophages treated with DVDMS-SDT are evaluated by RNA sequencing, and the rich gene biological processes are analyzed, and the changes are identified in lipid metaboly-related pathways significantly. (C-D) Westernblot representatives and statistical graphs of diacylglycerol kinase E (DGKE) in M1 macrophages, M1 macrophages treated with DVDMS-SDT, M1 macrophages +Rankl, M1 macrophages treated with DVDMS-SDT+Rankl, M1 macrophages treated with DVDMS-SDT +Rankl+NAC. n = 6. *P < 0.05. (E) DGKE mRNA expressions in M1 macrophages, M1 macrophages treated with DVDMS-SDT, M1 macrophages treated with DVDMS-SDT +NAC, and M1 macrophages treated with DVDMS-SDT +R59022. n = 3. *P < 0.05, **P < 0.01. (F) Determination of PA content of macrophages in M2 macrophages, M1 macrophages, different time after treatment of M1 macrophages by DVDMS-SDT, M1 macrophages treated with DVDMS-SDT +NAC. n = 3. *P < 0.05, **P < 0.01, ****P < 0.0001. (G)Determination of PA content of macrophages in M1 macrophages, M1 macrophages treated with RANKL, M1 macrophages treated with DVDMS-SDT+RANKL, M1 macrophages treated with DVDMS-SDT+RANKL+NAC, M1 macrophages treated with DVDMS-SDT+RANKL+R59022. (H) Alizarin red staining representatives and statistical graphs of direct co-culture system of calcified MOVAS cells and M1 macrophages, M1 macrophages treated with SDT, M1 macrophages treated with SDT+NAC, M1 macrophages treated with PA. Scale =200um. n = 3. *P < 0.05, **P < 0.01.

Western blot and RT-PCR showed that the expressions of DGKE protein and mRNA in M1 macrophages treated with SDT were significantly up-regulated. In the presence of Rankl, SDT further increased the expression of DGKE protein and mRNA in M1 macrophages. The effect of SDT was inhibited by ROS scavenger N-acetylcysteine (NAC). Our results indicated that SDT enhanced the expression of PA synthetase DGKE through ROS (Figure 6C-E). Elisa showed that SDT increased the PA expression of M1 macrophages. In the presence of Rankl, SDT further increased the PA expression of M1 macrophages. The PA expression of M1 macrophages was significantly increased from 3 hours after SDT, and the effect lasted for at least 24 hours. The PA expression of M1 macrophages treated with SDT was inhibited by NAC and DGKs inhibitor R59022. Our results indicated that SDT promoted the expression of DGKE and PA in M1 macrophages through ROS (Figure 6F,G). The Alizarin red staining showed that SDT reduced the degree of calcification of direct co-culture system of M1 macrophages and calcified MOVAS cells through ROS, while M1 macrophages treated with exogenous PA had a similar effect as SDT (Figure 6H-J).

### SDT promotes macrophage fusion, enhances CA2 expression and alleviates MOVAS cells calcification through ROS-DGKE-PA pathway

To explore whether SDT could stimulate M1 macrophages to fuse, atomic force microscopy (AFM) was applied and the results showed the diameter of M1 macrophages treated with SDT increased from about 10um to more than 30um, indicating that the M1 macrophages had fused (Figure 7A,B). To explore whether SDT promotes M1 macrophage to fuse by ROS-DGKE-PA pathway, phalloidin and DAPI were used to stain the actin and nucleu in M1 macrophages respectively. Our results showed that M1 macrophages treated with SDT had a higher degree of fusion and more nucleus. This fusion could be inhibited by NAC and DGKs inhibitor R59022, and could be restored by exogenous PA (Figure 7C,E). When the duration of SDT was 7 minutes, the typical multinucleated osteoclast-like cells appeared at 24th hours after SDT (Figure 7D,F). Tartrate resistant acid phosphatase (TRAP) staining showed that the number of TRAP-positive M1 macrophages increased after SDT (Figure S7G-H). Our results indicated that SDT promoted the fusion of M1 macrophages into typical multinucleated osteoclast-like cells through ROS-DGKE-PA pathway.

**Figure 7.**
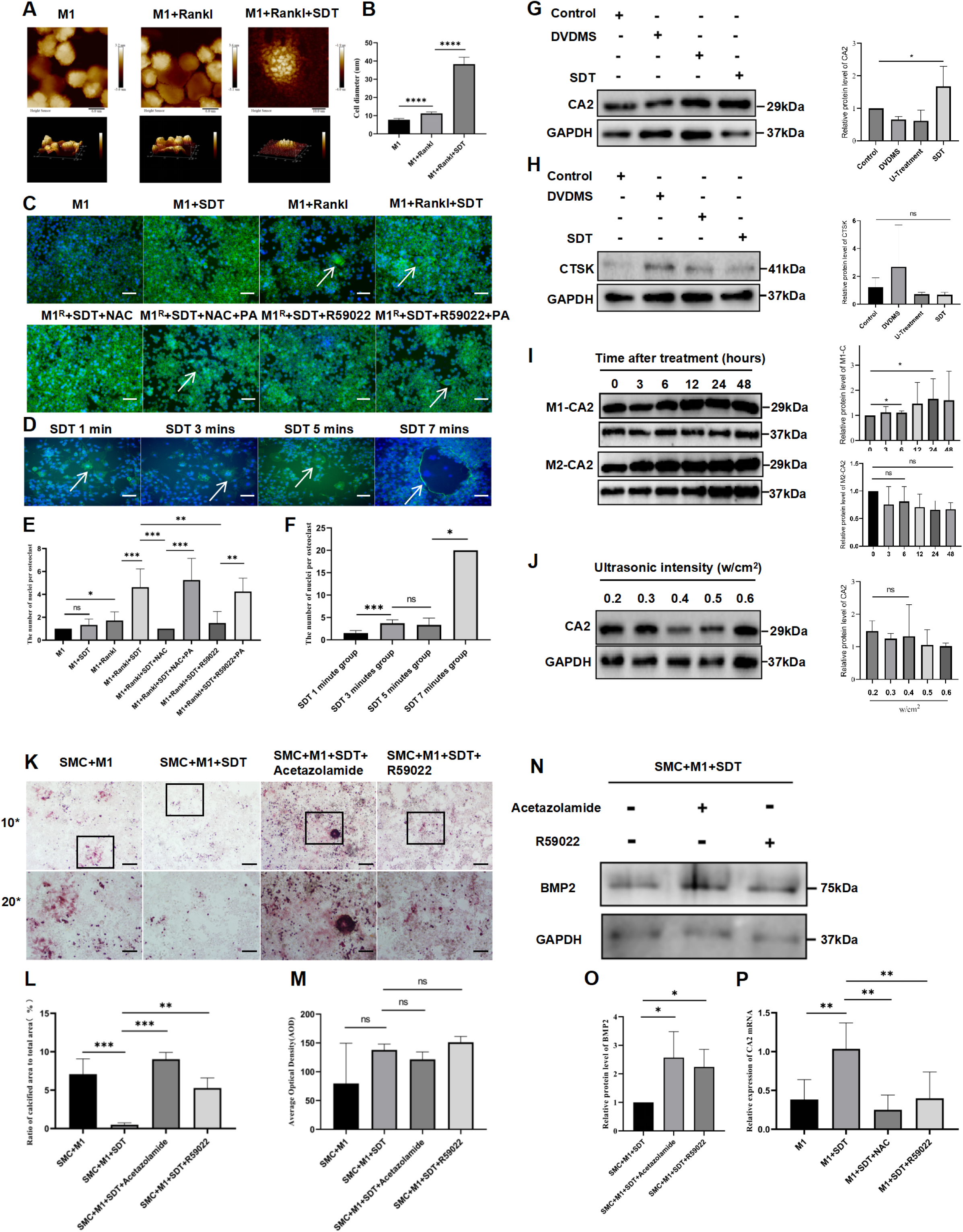
SDT promotes macrophage fusion, enhances CA2 expression and alleviates MOVAS cells calcification through ROS-DGKE-PA pathway. (A-B) AFM representatives and statistical graphs of M1 macrophages, M1 macrophages treated with RANKL, M1 macrophages treated with SDT+RANKL. n = 3. ****P < 0.0001. (C,E) Representative of phalloidin staining actin and DAPI staining nucleus of M1 macrophages, M1 macrophages treated with SDT, M1 macrophages treated with RANKL, M1 macrophages treated with SDT+RANKL, M1 macrophages treated with SDT+RANKL+NAC, M1 macrophages treated with SDT+RANKL+NAC+PA,.M1 macrophages treated with SDT+RANKL+R59022, M1 macrophages treated with SDT+RANKL+R59022+PA. Scale =100um. (D,F) Representative of phalloidin staining actin and DAPI staining nucleus of M1 macrophages undergoing different SDT durations. (G) Westernblot representatives and statistical graphs of M1 macrophage CA2 in control group, only DVDMS group, only ultrasound group, SDT group. n = 3. **P < 0.01. (H) Westernblot representatives and statistical graphs of M1 macrophage CTSK expression in control group, only DVDMS group, only ultrasound group, SDT group. n = 3. (I) Westernblot representatives and statistical graphs of CA2 in M1 macrophages or M2 macrophages at 0h, 3h, 6h, 12h, 24h, 48h after SDT. n = 3. *P < 0.05. (J) Westernblot representatives and statistical graphs of CA2 in M1 macrophages treated with 0.2w/cm^2^, 0.3w/cm^2^, 0.4w/cm^2^, 0.5w/cm^2^, 0.6w/cm^2^ ultrasonic intensity of SDT. n = 3. *P < 0.05. (K-M) Alizarin red staining representatives and statistical graphs of direct co-culture system of calcified MOVAS cells and M1 macrophages, M1 macrophages treated with SDT, M1 macrophages treated with SDT+acetazolamide, M1 macrophages treated with SDT+R59022, respectively. Scale =500um. n = 3. *P < 0.05, **P < 0.01. 10* scale =200um,20* scale =100um. (N-O) Westernblot representatives and statistical graphs of BMP2 expression in direct co-culture system of calcified MOVAS cells and M1 macrophages treated with SDT, M1 macrophages treated with SDT+acetazolamide, M1 macrophages treated with SDT+R59022, respectively. n = 4. *P < 0.05. (P) mRNA expression of CA2 in M1 macrophages, M1 macrophages treated with SDT, M1 macrophages treated with SDT+NAC, M1 macrophages treated with SDT+R59022. n = 3. **P < 0.01.

To explore whether SDT could enhance osteoclast function of M1 macrophages, westernblot and RT-PCR were used to detect the expression of CA2 and cathepsin K (CTSK), the key osteoclast enzymes. Our results showed that SDT up-regulated the expression of CA2 significantly, while SDT did not change the expression of CTSK significantly (Figure 7G,H). To examine the optimal duration and intensity of SDT, westernblot was used and the results showed that SDT up-regulated the CA2 expression significantly 6 hours after SDT, while there was no significant difference in CA2 expression among different intensity of SDT within the range of 0.2w/cm^2^ to 0.6w/cm^2^ (Figure 7I,J). Compared with M1 macrophages, M2 macrophages expressed CA2 highly (Figure S6E). This suggested that SDT could increase CA2 expression but not CTSK in M1 macrophages.

To explore whether SDT could reduce MOVAS cells calcification through ROS-DGKE-PA pathway of M1 macrophages, Alizarin red staining and westernblot were used and the results showed that SDT reduced the degree of calcification of direct co-culture system of M1 macrophages and MOVAS cells. DGKs inhibitor R59022 and CA2 inhibitors acetazolamide could reduce the effects of SDT on M1 macrophages and further indicated that SDT could reduce MOVAS cells calcification through ROS-DGKE-PA pathway of M1 macrophages (Figure 7K-P). To explore whether SDT could enhance the decalcification function of M1 macrophages, the inorganic model of microcalcification was applied. The results showed that M1 macrophages treated with SDT reduced the area of microcalcification and increased the average optical density of microcalcification. SDT did not change the bone destructive ability of M1 macrophages on bovine bone slices significantly (Figure S7A-E). The calcium content in the direct co-culture system of M1 macrophages or M2 macrophages and calcified MOVAS cells decreased after SDT (Figure S7F). These results revealed that SDT could enhance the decalcification function.

### SDT alleviated early calcification by increasing CA2 expression in macrophages in early calcified areas of ApoE-/- mice

To verify whether SDT could increase CA2 expression in macrophages in early calcified areas of ApoE-/- mice, immunofluorescence staining assay was used. Our results showed that CA2 expressed by macrophages and the calcification related protein BMP2 were located in the same region (Figure S8A-C). CA2 expression in macrophages in early calcified areas of ApoE-/- mice increased at the 15th day after SDT, but did not increase at the 30th day after SDT (Figure 8A,B). To explore whether SDT alleviated early calcification by increasing CA2 expression in macrophages in early calcified areas of ApoE-/- mice, acetazolamide was used to inhibit the expression of CA2 in ApoE-/- mice. The results showed that acetazolamide down-regulated the expression of CA2 and up-regulated the expressions of BMP2 and OST respectively. Acetazolamide reversed the effects of SDT on the expressions of CA2 and BMP2, OST individually (Figure 8C,D). The results demonstrated that SDT alleviated early calcification by increasing CA2 expression in macrophages in early calcified areas of ApoE-/- mice.

**Figure 8.**
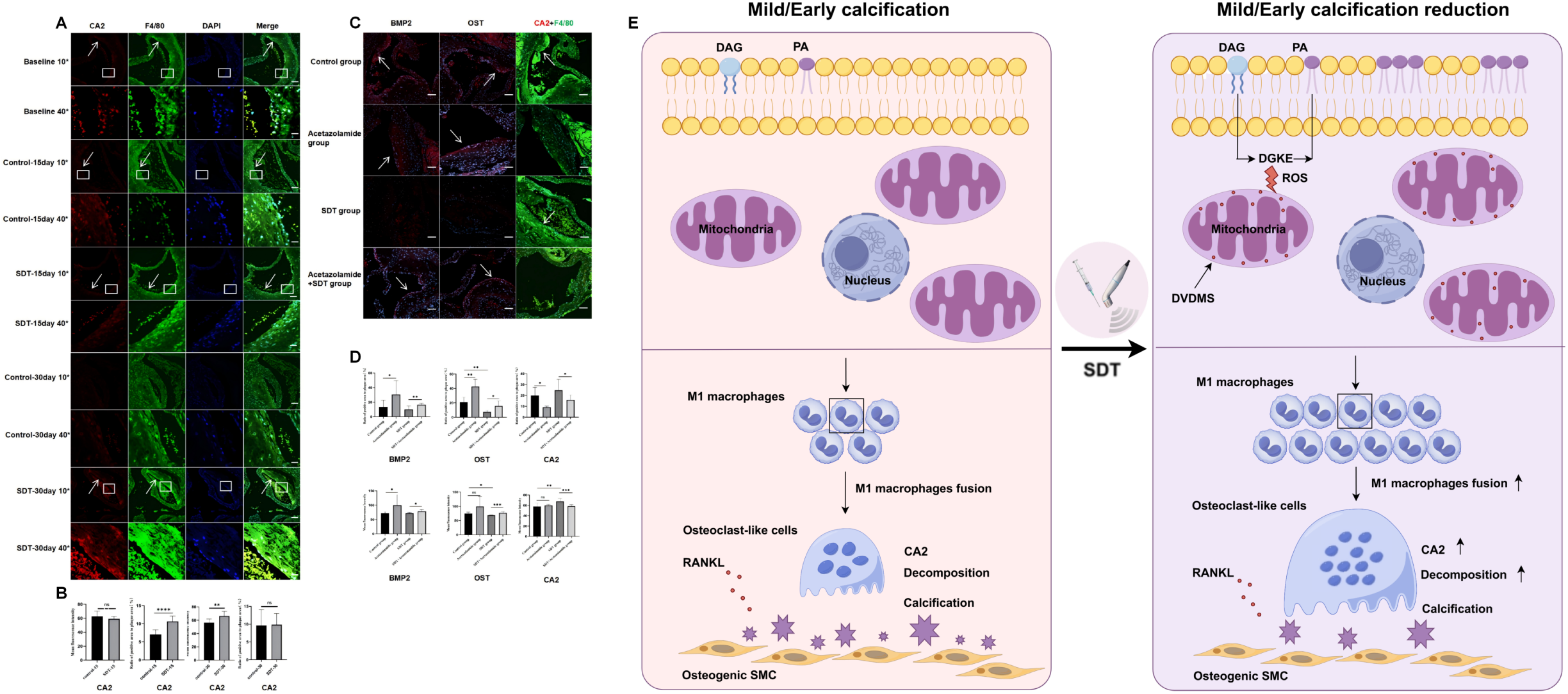
SDT alleviates plaque calcification by increasing CA2 expression in macrophages in early calcified areas of ApoE-/- mice. (A) Immunofluorescence staining representatives of CA2 in early calcification model of atherosclerotic plaques in ApoE-/- mice before and after SDT (10* scale =200um, 40* scale =50um). (B) The percentage of CA2 positive region in the total plaque area and the mean fluorescence intensity. n = 10 in the control-15day group, n = 7 in the SDT-15day group, n = 9 in the control-30day group, and n = 9 in the SDT-30day group. (C) Immunofluorescence staining representatives of F4/80, calcification related proteins BMP2, OST, and CA2 of macrophages with atherosclerotic plaques in early calcification model of ApoE-/- mice in control group, acetazolamide group, SDT group, acetazolamide+SDT group. ( scale =200um). (D) Statistical graphs of calcification related proteins BMP2, OST, and CA2 of macrophages in early calcified plaques of ApoE-/- mice in control group, acetazolamide group, SDT group, acetazolamide+SDT group. BMP2-control group n = 9, BMP2-acetazolamide group n = 9, BMP2-SDT group n = 9, BMP2-SDT+acetazolamide group n = 9. OST-Control group n = 6, OST-acetazolamide group n = 4,OST-SDT group n = 5, OST-SDT+acetazolamide group n = 6. CA2-control group n = 4, CA2-acetazolamide group n = 4, CA2-SDT group n = 6, and CA2-SDT+acetazolamide group n = 8. (scale =200um). (E) Mechanism of SDT alleviating Mild/Early calcification. CA2 indicates Carbonic anhydrase 2; RANKL, Receptor Activator of Nuclear Factor-κ B Ligand; SMC, Smooth muscle cells; DAG, Diacylglycerol; PA, Phosphatidic acid; DGKE, Diacylglycerol kinase Ε; ROS, Reactive oxygen; DVDMS, Sinoporphyrin sodium; SDT, Sonodynamic therapy.

## DISCUSSION

The contributions of our study are as follows. SDT reduced mild calcification in patients with femoropopliteal PAD and early calcification in aortic atherosclerotic plaque in ApoE-/- mice. MSI revealed that SDT significantly increased the PA content in the early calcified region of ApoE-/- mice. Mendelian randomized analysis of single cell sequencing indicated that DGKE negatively regulated coronary artery calcification. PA mediated SDT facilitated M1 macrophage fusion and CA2 expression to improve the osteoclastic function and alleviate early calcification through ROS-DGKE-PA pathway (Figure 8E).

Currently, the number of calcified sites on PET/CT is widely used as a valid indicator for evaluating calcification load^36–38^. According to the Hounsfield unit calcium density of PET/CT, the lesions were divided into 3 groups: non-calcified (< 130 HU), mild calcified (130-399 HU) and severe calcified (≥400 HU)^39^. In our clinical trial, we adhered to this classification standard to establish groups and determine the number of calcification sites as the endpoint of exploration. Interestingly, the Multi-Ethnic Study of Atherosclerosis (MESA) demonstrated an independent association between biological age and calcification density, revealing that the natural progression of atherosclerosis corresponded to calcification density, with early calcification linked to low-density calcification^6^. Importantly, in our clinical trial, SDT significantly reduced the number of mildly calcified sites, the percentage of mildly calcified sites among total sites, and the target-to-background ratio (TBR) value of mildly calcified sites at the 30th day after SDT. Postmortem analysis indicated no correlation between the changes in the number of mildly calcified sites and the TBR value of mildly calcified sites before and after SDT. This suggests that the reduction in the number of mildly calcified sites due to SDT may not be dependent on inflammation reduction. Conversely, SDT did not significantly reduce the number of severely calcified sites, the percentage of severely calcified sites among total sites, or the TBR value of severely calcified sites at the 30th day after SDT. In fact, patients with stable coronary artery disease exhibited a high calcification density of atherosclerotic plaques^40–42^. Our clinical trial demostrated that SDT was effective only on mild calcification, and the effect was not depended on the reduction of inflammation.

In previous study, ApoE-/-mice were injected with two calcium tracers, and the aortic plaque microcalcification occured after 20 weeks high-fat diet, and then microcalcification fused to visible macrocalcification after 23 weeks high-fat diet^43^. This suggested that persistent high-fat diet might promote early microcalcification as a precursor to the formation of macrocalcification^43^. In our animal study, the 20th week of high-fat diet was set as the early calcification time point of ApoE-/- mice. And at this time point, SDT was carried out. At the 15th day and 30th day after SDT, the samples was taken. Immunofluorescence and immunohistochemistry showed that the calcified area of ApoE-/- mice were significantly reduced at the 15th day after SDT; however, it were not significantly reduced at the 30th day after SDT. This results might be related to the formation of macrocalcification. Micro-CT also showed that the calcified volume of ApoE-/- mice at the 15th day after SDT were significantly reduced. Our results from animal study further demostrated that SDT was mainly effective on early or mild calcification.

MSI was applied to identify the properties, quantitative variations, and spatial distributions of a variety of material molecules in a sample without labeling^44–46^. In previous study, MSI was conducted on human carotid atherosclerotic specimens and 55 types of lipids were identified to exhibit characteristic spatial distribution^47^. Our study was the first to apply MSI to find that SDT could significantly increase PA content in early calcification area of ApoE-/- mice. PA, 1, 2-diacyl-glycerol-3 phosphate, is one of the simplest membrane phospholipids and is present in almost all cell membranes^19^. PA is produced by three main ways in the cell: phospholipase D (PLD) hydrolyzed phosphatidylcholine; Diacylglycerol (DAG) is phosphorylated by diacylglycerol kinases (DGKs); Lysophosphatidyl acyltransferase (LPAATs)^19^. The key features of PA metabolism are redundancy and multilevel regulation, combined with the ability of PA biosynthesis to partially compensate each other, making the study of the biological function of specific PA pools challenging^32^. Our important finding indicated that PA might play a pivotal role in the reduction of early calcification.

Mendelian Randomization analysis is an important method to assess the causal relationship between exposure (e.g., lifestyle, cytokines, etc.) and outcomes (e.g., disease or health condition). Mendelian randomization analysis was performed to found a causal relationship between coronary artery calcification and acute inferior myocardial infarction^48^. Spatial transcriptomics combined with Mendelian randomization analysis was applied to find a causal relationship between high circulating levels of MMP9 and risk of atherosclerosis^49^. Our study was the first to combine single-cell sequencing Mendelian randomization analysis with MSI to explore the causal relationship between specific PA pools and vascular calcification. And we found that the PA pool derived from DGKE negatively regulated vascular calcification in monocyte/macrophages. In mammals, there are 10 known subtypes of DGKs, and DGKE is the only DGK subtype permanently attached to cell membranes. DGKE is involved in the regulation of many diseases^50^. Previous studies had shown that inhibiting DGKs could reduce ROS production in RAW264.7 macrophages^51^. Considering the results of our clinical trial and animal study, we speculated that SDT may negatively regulate vascular calcification through DGKE-PA.

We identified both M1 and M2 macrophages in the early calcification region of ApoE-/- mice. This finding raised an intriguing question: which macrophage subtype-M1 or M2 plays a predominant role in alleviating MOVAS cell calcification after SDT? In our study, direct co-culture of M1 or M2 macrophages with MOVAS cells was conducted respectively. And we found that, the co-culture system of M2 macrophages and MOVAS cells had a lower degree of calcification; whereas, the co-culture system of M1 macrophages and MOVAS cells had a higher degree of calcification. We further found that, SDT did not reduce the degree of calcification in the co-culture system of M2 macrophages and MOVAS cells significantly. Whereas, SDT reduced significantly the degree of calcification in the co-culture system of M1 macrophages and MOVAS cells. The effect of SDT depended on the contact between M1 macrophages and MOVAS cells. In a study involving 687 patients undergoing advanced carotid endarterectomy (CEA), plaques with microcalcification were associated with a high presence of M1 macrophages, while plaques with macrocalcification were predominantly composed of M2 macrophages^52^. Our in vitro study indicated that M1 macrophages played a major role in alleviating MOVAS cells calcification after SDT, with this effect being dependent on the direct contact between M1 macrophages and MOVAS cells.

In our clinical trial, postmortem analysis suggested that the reduction in the number of mildly calcified sites after SDT might not depend on the reduction of inflammation. This implied that SDT might have other underlying mechanisms. In our animal study, we applied MSI to reveal that SDT significantly increased PA content in early calcification area of ApoE-/- mice. PA promoted the fusion of macrophages and enhance osteoclastic function^20^. In our Mendelian randomization analysis, we found that the PA pool derived from DGKE negatively regulated vascular calcification in monocyte/macrophages. In our vitro study, we verified that the effect of SDT depended on the contact between M1 macrophages and MOVAS cells. Osteoclastic function required the contact between osteoclasts and calcification^53–55^. Based on the above results, we hypothesized that SDT alleviated early calcification by promoting M1 macrophage fusion and the osteoclastic function through DGKE-PA pathway. To verify this hypothesis, we used the osteoclast agonist Rankl to M1 macrophages and found that Rankl significantly increased the PA content of M1 macrophages. SDT further increased the PA content of M1 macrophages on the basis of Rankl. The effect of SDT on M1 macrophage PA could be inhibited by NAC or DGKE inhibitor R59022. The effect of SDT on MOVAS cells calcification could also be inhibited by NAC or DGKE inhibitor R59022. The effect of SDT on MOVAS cells calcification was restored by exogenous PA. These results demonstrated that SDT alleviated MOVAS cells calcification via ROS-DGKE-PA pathway.

Importantly, osteoclasts were large polynucleated cells that constructed a closed bone absorption microenvironment on the surface of the mineralized matrix and released hydrogen ions into the absorption cavity through the action of CA2 (H2O + CO2→HCO3^-^ + H^+^), acidifying and decomposing hydroxyapatite into Ca2^+^, H3PO4, H2CO3, water, and other substances^53–55^. CA2 knockout mice exhibited extensive vascular calcification^56^. Increased macrophage fusion significantly increased CA2 expression in osteoclasts^57,58^. Decreased macrophage fusion significantly decreased CA2 expression in osteoclasts^59^. In addition, osteoclasts released several hydrolases, such as CTSK and matrix metalloproteinase (MMP) members, to digest the organic components of the matrix, such as type I collagen^53–55^. In vitro calcification study found that collagen inhibited early microcalcification during the formation of early microcalcification^42^. Early calcification was dominated by hydroxyapatite^60^. Therefore, CA2 rather than CTSK may be more important for the treatment of early calcification. In this study, AFM showed that the diameter of M1 macrophages in the SDT group was significantly increased, indicating that M1 macrophages fusion occured. Immunofluorescence assay showed that M1 macrophages in SDT group had higher fusion degree and osteoclast-like cells had more nucleus. This fusion could be inhibited by NAC and DGKs inhibitor R59022, and could be restored by exogenous PA. After SDT, M1 macrophage CA2 was significantly up-regulated, but CTSK did not change significantly. SDT could reduce the calcification degree in direct co-culture system of M1 macrophages and calcified MOVAS cells. And the therapeutic effect of SDT could be inhibited by R59022 and CA2 inhibitor acetazolamide. Therefore, SDT facilitated M1 macrophage fusion and CA2 expression to improve the osteoclastic function and alleviate early calcification through ROS-DGKE-PA pathway.

In the present animal study, immunofluorescence assay and correlation analysis showed that M1 macrophage CA2 was spatially in the same region as the calcification related protein BMP2. This verified that SDT reduced early calcification through the contact of M1 macrophage CA2 and early calcification related protein BMP2. The animal result was correspond to the result of our in vitro study. On the 15th day after SDT, the percentage of BMP2-positive area decreased and the percentage of CA2-positive area of macrophages increased. On the 30th day after SDT, the percentage of BMP2-positive area and the percentage of CA2-positive area of macrophages had no significant change. The effect of SDT on CA2 and the calcification related protein BMP2, OST could be inhibited by acetazolamide. This animal study also demostrated that SDT alleviated early calcification through M1 macrophage CA2.

Presently, calcification therapy targeting osteoclastic macrophages faces many safety challenges^15^. The resonant frequency of ultrasonic transducer applied in this study is 1.0 MHz and the ultrasound intensity of ultrasonic transducer is 1.8 W/cm2. The ultrasound can not penetrate the bone and affect osteoclast activity^61–64^. In the present in vitro study, flow cytometry revealed that macrophages effectively phagocytosed DVDMS, whereas MOVAS cells did not phagocytose DVDMS after 4 hours of individual incubation. Our previous animal study indicated that SDT utilized the different absorption capacities of various cells in plaques to target macrophages while minimizing interference with smooth muscle cells, endothelial cells, and other cell types^27,65^. Thus, PA-mediated SDT has the potential to address these safety challenges effectively.

### Conclusions

In conclusion, PA mediated SDT facilitates M1 macrophage fusion and CA2 expression to improve the osteoclastic function and alleviate early calcification through ROS-DGKE-PA pathway. SDT is a novel and promising strategy for early vascular calcification.

## Supplemental Material

Expanded Methods

Figures S1-S8

References 66-75

## Data Availability

Additional data are made available in supplementary materials of this manuscript. The authors will supply the relevant data in response to reasonable requests.

https://www.clinicaltrials.gov

## Nonstandard Abbreviations and Acronyms

SDT: sonodynamic therapy
PAD: peripheral artery disease
PET/CT: positron emission tomography/computed tomography
TBR: target-to-background ratio
DGKE: diacylglycerol kinase E
PA: phosphatidic acid
Rankl: nuclear factor κB receptor activating factor ligand
MOVAS: cells mouse aortic vascular smooth muscle cells
ROS: reactive oxygen species
CA2: carbonic anhydroase II
MSI: Mass spectrometry imaging
DVDMS: Sinoporphyrin sodium
BMP2: bone morphogenetic protein 2
OST: osteocalcin
RUNX2: osteogenic transcription factor
OPN: osteopontin
NAC: N-acetylcysteine

## REFERENCES

1. Hutcheson Joshua D, Goettsch Claudia, Cardiovascular Calcification Heterogeneity in Chronic Kidney Disease. [J]. Circ Res, 2023, 132: 993–1012.

2. Demer Linda L, Tintut Yin, Interactive and Multifactorial Mechanisms of Calcific Vascular and Valvular Disease. [J]. Trends Endocrinol Metab, 2019, 30: 646–657.

3. Onnis Carlotta, Virmani Renu, Kawai Kenji, et al. Coronary Artery Calcification: Current Concepts and Clinical Implications. [J]. Circulation, 2024, 149: 251–266.

4. Nakahara Takehiro, Dweck Marc R, Narula Navneet, et al. Coronary Artery Calcification: From Mechanism to Molecular Imaging. [J]. JACC Cardiovasc Imaging, 2017, 10: 582–593.

5. Rogers Maximillian A, Aikawa Elena, Cardiovascular calcification: artificial intelligence and big data accelerate mechanistic discovery. [J]. Nat Rev Cardiol, 2019, 16: 261–274.

6. Criqui Michael H, Denenberg Julie O, Ix Joachim H, et al. Calcium density of coronary artery plaque and risk of incident cardiovascular events. [J]. JAMA, 2014, 311: 271–8.

7. Criqui Michael H, Knox Jessica B, Denenberg Julie O, et al. Coronary Artery Calcium Volume and Density: Potential Interactions and Overall Predictive Value: The Multi-Ethnic Study of Atherosclerosis. [J]. JACC Cardiovasc Imaging, 2017, 10: 845–854.

8. Razavi Alexander C, Agatston Arthur S, Shaw Leslee J, et al. Evolving Role of Calcium Density in Coronary Artery Calcium Scoring and Atherosclerotic Cardiovascular Disease Risk. [J]. JACC Cardiovasc Imaging, 2022, 15: 1648–1662.

9. Cowell SJ, Newby DE, Prescott RJ, Bloomfield P, Reid J, Northridge DB, Boon NA; Scottish Aortic Stenosis and Lipid Lowering Trial, Impact on Regression (SALTIRE) Investigators. A randomized trial of intensive lipid-lowering therapy in calcific aortic stenosis. N Engl J Med. 2005 Jun 9;352(23):2389–97.

10. Puri R, Nicholls SJ, Shao M, Kataoka Y, Uno K, Kapadia SR, Tuzcu EM, Nissen SE. Impact of statins on serial coronary calcification during atheroma progression and regression. J Am Coll Cardiol. 2015 Apr 7;65(13):1273–1282.

11. Gomez D, Baylis RA, Durgin BG, Newman AAC, Alencar GF, Mahan S, St Hilaire C, Müller W, Waisman A, Francis SE, Pinteaux E, Randolph GJ, Gram H, Owens GK. Interleukin-1β has atheroprotective effects in advanced atherosclerotic lesions of mice. Nat Med. 2018 Sep;24(9):1418–1429.

12. Ouyang Liu, Yu Changjiang, Xie Zhiyong, et al. Indoleamine 2,3-Dioxygenase 1 Deletion-Mediated Kynurenine Insufficiency in Vascular Smooth Muscle Cells Exacerbates Arterial Calcification. [J]. Circulation, 2022, 145: 1784–1798.

13. Wu Meiting, Rementer Cameron,Giachelli Cecilia M, Vascular calcification: an update on mechanisms and challenges in treatment. [J]. Calcif Tissue Int, 2013, 93: 365–73.

14. Qiao JH, Mishra V, Fishbein MC, Sinha SK, Rajavashisth TB. Multinucleated giant cells in atherosclerotic plaques of human carotid arteries: Identification of osteoclast-like cells and their specific proteins in artery wall. Exp Mol Pathol. 2015 Dec;99(3):654–62.

15. Rogers Maximillian A, Aikawa Masanori, Aikawa Elena, Macrophage Heterogeneity Complicates Reversal of Calcification in Cardiovascular Tissues. [J]. Circ Res, 2017, 121: 5–7.

16. Chinetti-Gbaguidi Giulia, Daoudi Mehdi, Rosa Mickael, et al. Human Alternative Macrophages Populate Calcified Areas of Atherosclerotic Lesions and Display Impaired RANKL-Induced Osteoclastic Bone Resorption Activity. [J]. Circ Res, 2017, 121: 19–30.

17. Canet-Soulas Emmanuelle, Bessueille Laurence,Mechtouff Laura, et al. The Elusive Origin of Atherosclerotic Plaque Calcification. [J]. Front Cell Dev Biol, 2021, 9: 622736.

18. Vickers Kasey C, Castro-Chavez Fernando,Morrisett Joel D, Lyso-phosphatidylcholine induces osteogenic gene expression and phenotype in vascular smooth muscle cells. [J]. Atherosclerosis, 2010, 211: 122–9.

19. Zhukovsky MA, Filograna A, Luini A, Corda D, Valente C. Phosphatidic acid in membrane rearrangements. FEBS Lett. 2019 Sep;593(17):2428–2451.

20. Park MK, Her YM, Cho ML, Oh HJ, Park EM, Kwok SK, Ju JH, Park KS, Min DS, Kim HY, Park SH. IL-15 promotes osteoclastogenesis via the PLD pathway in rheumatoid arthritis. Immunol Lett. 2011 Sep 30;139(1-2):42–51.

21. Tappia PS, Dent MR, Dhalla NS. Oxidative stress and redox regulation of phospholipase D in myocardial disease. Free Radic Biol Med. 2006 Aug 1;41(3):349–61.

22. Zhang Yi, Zhang Xiangqian,Yang Huocheng, et al. Advanced biotechnology-assisted precise sonodynamic therapy. [J]. Chem Soc Rev, 2021, 50: 11227–11248.

23. Zhang Chi, Pu Kanyi, Organic Sonodynamic Materials for Combination Cancer Immunotherapy. [J]. Adv Mater, 2023, 35: e2303059.

24. Wang Huan, Yang Yang,Sun Xin, et al. Sonodynamic therapy-induced foam cells apoptosis activates the phagocytic PPARγ-LXRα-ABCA1/ABCG1 pathway and promotes cholesterol efflux in advanced plaque. [J]. Theranostics, 2018, 8: 4969–4984.

25. Yang Yang, Wang Jiayu, Guo Shuyuan, et al. Non-lethal sonodynamic therapy facilitates the M1-to-M2 transition in advanced atherosclerotic plaques via activating the ROS-AMPK-mTORC1-autophagy pathway. [J]. Redox Biol, 2020, 32: 101501.

26. Sun X, Guo SY, Yao JT, Wang H, Peng CH, Li BC, Wang Y, Jiang YX, Wang TY, Yang Y, Cheng JL,Wang W, Cao ZY, Cao, WW, Li M, Tian Y. Rapid inhibition of atherosclerotic plaque progression by sonodynamic therapy, Cardiovas Res. 2019, 15(1):190–203.

27. Yao J, Gao W, Wang Y, et al. Sonodynamic Therapy Suppresses Neovascularization in Atherosclerotic Plaques via Macrophage Apoptosis-Induced Endothelial Cell Apoptosis. JACC: Basic to Translational Science. 2020; 5(1):53–65.

28. Jiang Yongxing, Fan Jingxue, Li Yong, et al. Rapid reduction in plaque inflammation by sonodynamic therapy inpatients with symptomatic femoropopliteal peripheral artery disease:A randomized controlled trial. [J]. Int J Cardiol, 2021, 325: 132–139.

29. Wang Y, Wang W, Xu H, et al. Non-lethal sonodynamic therapy inhibits atherosclerotic plaque progression in ApoE−/−mice and attenuates ox-LDL-mediated macrophage impairment by inducing hemeoxygenase-1. Cell Physiol Biochem. 2017;41(6):2432–2446.

30. Yan J, Horng T. Lipid Metabolism in Regulation of Macrophage Functions. Trends Cell Biol. 2020 Dec;30(12):979–989.

31. Vassiliou E, Farias-Pereira R. Impact of Lipid Metabolism on Macrophage Polarization: Implications for Inflammation and Tumor Immunity. Int J Mol Sci. 2023 Jul 27;24(15):12032.

32. Tei R, Baskin JM. Spatiotemporal control of phosphatidic acid signaling with optogenetic, engineered phospholipase Ds. J Cell Biol. 2020 Mar 2;219(3):e201907013.

33. Ding R, Wang Q, Gong L, Zhang T, Zou X, Xiong K, Liao Q, Plass M, Li L. scQTLbase: an integrated human single-cell eQTL database. Nucleic Acids Res. 2024 Jan 5;52(D1):D1010–D1017.

34. Passos LSA, Lupieri A, Becker-Greene D, Aikawa E. Innate and adaptive immunity in cardiovascular calcification. Atherosclerosis. 2020 Aug;306:59–67.

35. New SE, Aikawa E. Molecular imaging insights into early inflammatory stages of arterial and aortic valve calcification. Circ Res. 2011 May 27;108(11):1381–91.

36. Derlin T, Thiele J, Weiberg D, Thackeray JT, Püschel K, Wester HJ, Aguirre Dávila L, Larena-Avellaneda A, Daum G, Bengel FM, Schumacher U. Evaluation of 68Ga-Glutamate Carboxypeptidase II Ligand Positron Emission Tomography for Clinical Molecular Imaging of Atherosclerotic Plaque Neovascularization. Arterioscler Thromb Vasc Biol. 2016 Nov;36(11):2213–2219.

37. Weiberg D, Thackeray JT, Daum G, Sohns JM, Kropf S, Wester HJ, Ross TL, Bengel FM, Derlin T. Clinical Molecular Imaging of Chemokine Receptor CXCR4 Expression in Atherosclerotic Plaque Using 68Ga-Pentixafor PET: Correlation with Cardiovascular Risk Factors and Calcified Plaque Burden. J Nucl Med. 2018 Feb;59(2):266–272.

38. Kosmala A, Serfling SE, Michalski K, Lindner T, Schirbel A, Higuchi T, Hartrampf PE, Derlin T, Buck AK, Weich A, Werner RA. Molecular imaging of arterial fibroblast activation protein: association with calcified plaque burden and cardiovascular risk factors. Eur J Nucl Med Mol Imaging. 2023 Aug;50(10):3011–3021.

39. Li X, Heber D, Cal-Gonzalez J, Karanikas G, Mayerhoefer ME, Rasul S, Beitzke D, Zhang X, Agis H, Mitterhauser M, Wadsak W, Beyer T, Loewe C, Hacker M. Association Between Osteogenesis and Inflammation During the Progression of Calcified Plaque Evaluated by 18F-Fluoride and 18F-FDG. J Nucl Med. 2017 Jun;58(6):968–974.

40. Schoenhagen P, Ziada KM, Kapadia SR, Crowe TD, Nissen SE, Tuzcu EM. Extent and direction of arterial remodeling in stable versus unstable coronary syndromes: an intravascular ultrasound study. Circulation. 2000 Feb 15;101(6):598–603.

41. Leber AW, Knez A, White CW, Becker A, von Ziegler F, Muehling O, Becker C, Reiser M, Steinbeck G, Boekstegers P. Composition of coronary atherosclerotic plaques in patients with acute myocardial infarction and stable angina pectoris determined by contrast-enhanced multislice computed tomography. Am J Cardiol. 2003 Mar 15;91(6):714–8.

42. Shemesh J, Apter S, Itzchak Y, Motro M. Coronary calcification compared in patients with acute versus in those with chronic coronary events by using dual-sector spiral CT. Radiology. 2003 Feb;226(2):483–8.

43. Hutcheson JD, Goettsch C, Bertazzo S, Maldonado N, Ruiz JL, Goh W, Yabusaki K, Faits T, Bouten C, Franck G, Quillard T, Libby P, Aikawa M, Weinbaum S, Aikawa E. Genesis and growth of extracellular-vesicle-derived microcalcification in atherosclerotic plaques. Nat Mater. 2016 Mar;15(3):335–43.

44. Harkewicz R, Dennis EA. Applications of mass spectrometry to lipids and membranes. Annu Rev Biochem. 2011;80:301–25.

45. Norris JL, Caprioli RM. Analysis of tissue specimens by matrix-assisted laser desorption/ionization imaging mass spectrometry in biological and clinical research. Chem Rev. 2013 Apr 10;113(4):2309–42.

46. Chughtai K, Heeren RM. Mass spectrometric imaging for biomedical tissue analysis. Chem Rev. 2010 May 12;110(5):3237–77.

47. Li W, Luo J, Peng F, Liu R, Bai X, Wang T, Zhang X, Zhu J, Li XY, Wang Z, Liu W, Wang J, Zhang L, Chen X, Xue T, Ding C, Wang C, Jiao L. Spatial metabolomics identifies lipid profiles of human carotid atherosclerosis. Atherosclerosis. 2023 Jan;364:20–28.

48. Sae-Jie W, Tangcharoen T, Vathesatogkit P, Aekplakorn W, Charoen P. Mendelian randomization study of the effect of coronary artery calcification on atherosclerotic cardiovascular diseases. Sci Rep. 2022 Sep 1;12(1):14829.

49. Sun J, Singh P, Shami A, Kluza E, Pan M, Djordjevic D, Michaelsen NB, Kennbäck C, van der Wel NN, Orho-Melander M, Nilsson J, Formentini I, Conde-Knape K, Lutgens E, Edsfeldt A, Gonçalves I. Spatial Transcriptional Mapping Reveals Site-Specific Pathways Underlying Human Atherosclerotic Plaque Rupture. J Am Coll Cardiol. 2023 Jun 13;81(23):2213–2227.

50. Jennings W, Doshi S, D’Souza K, Epand RM. Molecular properties of diacylglycerol kinase-epsilon in relation to function. Chem Phys Lipids. 2015 Nov;192:100–108.

51. Kamio N, Akifusa S, Yamashita Y. Diacylglycerol kinase alpha regulates globular adiponectin-induced reactive oxygen species. Free Radic Res. 2011 Mar;45(3):336–41.

52. Montanaro M, Scimeca M, Anemona L, Servadei F, Giacobbi E, Bonfiglio R, Bonanno E, Urbano N, Ippoliti A, Santeusanio G, Schillaci O, Mauriello A. The Paradox Effect of Calcification in Carotid Atherosclerosis: Microcalcification is Correlated with Plaque Instability. Int J Mol Sci. 2021 Jan 1;22(1):395.

53. Boyle William J, Simonet W Scott, Lacey David L, Osteoclast differentiation and activation. [J]. Nature, 2003, 423: 337–42.

54. Teitelbaum SL, Ross FP. Genetic regulation of osteoclast development and function. Nat Rev Genet. 2003 Aug;4(8):638–49.

55. Lacey DL, Boyle WJ, Simonet WS, Kostenuik PJ, Dougall WC, Sullivan JK, San Martin J, Dansey R. Bench to bedside: elucidation of the OPG-RANK-RANKL pathway and the development of denosumab. Nat Rev Drug Discov. 2012 May;11(5):401–19.

56. Spicer SS, Lewis SE, Tashian RE, Schulte BA. Mice carrying a CAR-2 null allele lack carbonic anhydrase II immunohistochemically and show vascular calcification. Am J Pathol. 1989 Apr;134(4):947–54.

57. Manrique E, Castillo LM, Lazala O, Guerrero CA, Acosta O. Bone resorptive activity of human peripheral blood mononuclear cells after fusion with polyethylene glycol. J Bone Miner Metab. 2017 Mar;35(2):127–141.

58. Xu XY, Guo C, Yan YX, Guo Y, Li RX, Song M, Zhang XZ. Differential effects of mechanical strain on osteoclastogenesis and osteoclast-related gene expression in RAW264.7 cells. Mol Med Rep. 2012 Aug;6(2):409–15.

59. Kim YH, Kim JL, Lee EJ, Park SH, Han SY, Kang SA, Kang YH. Fisetin antagonizes cell fusion, cytoskeletal organization and bone resorption in RANKL-differentiated murine macrophages. J Nutr Biochem. 2014 Mar;25(3):295–303.

60. Proudfoot, D.; Shanahan, C.M.; Weissberg, P.L. Vascular calcification: New insights into an old problem. J. Pathol. 1998, 185, 1–3.

61. Yang J, Gao P, Li Q, Wang T, Guo S, Zhang J, Zhang T, Wu G, Guo Y, Wang Z, Tian Y. Arterial Adventitial Vasa Vasorum Density Reflects The Progression Of Unstable Plaques: A Retrospective Clinical Study. Ultrasound Med Biol. 2024 May;50(5):712–721.

62. Bianchi S. Ultrasound and bone: a pictorial review. J Ultrasound. 2020 Sep;23(3):227-257. doi: 10.1007/s40477-020-00477-4. Epub 2020 May 17.

63. Hoffman DF, Adams E, Bianchi S. Ultrasonography of fractures in sports medicine. Br J Sports Med. 2015 Feb;49(3):152–60.

64. Javid A, Ilham S, Kiani M. A Review of Ultrasound Neuromodulation Technologies. IEEE Trans Biomed Circuits Syst. 2023 Oct;17(5):1084–1096.

65. Yao J, Zhao X, Tan F, Cao X, Guo S, Li X, Huang Z, Diabakte K, Wang L, Liu M, Shen Z, Li B, Cao Z, Sheng S, Lu M, Cao Y, Jin H, Zhang Z, Tian Y. Early modulation of macrophage ROS-PPARγ-NF-κB signalling by sonodynamic therapy attenuates neointimal hyperplasia in rabbits. Sci Rep. 2020 Jul 15;10(1):11638.

66. H. Emami, E. Vucic, S. Subramanian, A. Abdelbaky, Z.A. Fayad, S. Du, et al., The effect of BMS-582949, a P38 mitogen-activated protein kinase (P38 MAPK) inhibitor on arterial inflammation: a multicenter FDG-PET trial, Atherosclerosis. 240 (2) (2015) 490–496.

67. Brent MB, Simonsen U, Thomsen JS, Brüel A. Effect of Acetazolamide and Zoledronate on Simulated High Altitude-Induced Bone Loss. Front Endocrinol (Lausanne). 2022 Feb 9;13:831369.

68. Awan Z, Denis M, Bailey D, Giaid A, Prat A, Goltzman D, Seidah NG, Genest J. The LDLR deficient mouse as a model for aortic calcification and quantification by micro-computed tomography. Atherosclerosis. 2011 Dec;219(2):455–62.

69. Li P, Hao Z, Wu J, Ma C, Xu Y, Li J, Lan R, Zhu B, Ren P, Fan D, Sun S. Comparative Proteomic Analysis of Polarized Human THP-1 and Mouse RAW264.7 Macrophages. Front Immunol. 2021 Jun 29;12:700009.

70. Pang H, Xiao L, Lu Z, Chen H, Shang Z, Jiang N, Wang X, Wei F, Jiang A, Chen Y, Niu Y. Targeting androgen receptor in macrophages inhibits phosphate-induced vascular smooth muscle cell calcification by decreasing IL-6 expression. Vascul Pharmacol. 2020 Jul;130:106681.

71. Byon CH, Sun Y, Chen J, Yuan K, Mao X, Heath JM, Anderson PG, Tintut Y, Demer LL, Wang D, Chen Y. Runx2-upregulated receptor activator of nuclear factorκB ligand in calcifying smooth muscle cells promotes migration and osteoclastic differentiation of macrophages. Arterioscler Thromb Vasc Biol. 2011 Jun;31(6):1387–96.

72. Hudalla H, Michael Z, Christodoulou N, Willis GR, Fernandez-Gonzalez A, Filatava EJ, Dieffenbach P, Fredenburgh LE, Stearman RS, Geraci MW, Kourembanas S, Christou H. Carbonic Anhydrase Inhibition Ameliorates Inflammation and Experimental Pulmonary Hypertension. Am J Respir Cell Mol Biol. 2019 Oct;61(4):512–524.

73. Mackenzie NC, Zhu D, Longley L, Patterson CS, Kommareddy S, MacRae VE. MOVAS-1 cell line: a new in vitro model of vascular calcification. Int J Mol Med. 2011 May;27(5):663–8.

74. Barinda AJ, Ikeda K, Hirata KI, Emoto N. Macrophages Highly Express Carbonic Anhydrase 2 and Play a Significant Role in Demineralization of the Ectopic Calcification. Kobe J Med Sci. 2017 Oct 16;63(2):E45–E50.

75. Zhang P, Jiang H, Yang M, Bi C, Zhang K, Liu D, Wei M, Jiang Z, Lv K, Fang C, Liu J, Zhang T, Xu Y, Zhang J. AGK Potentiates Arterial Thrombosis by Affecting Talin-1 andαIIbβ3-Mediated Bidirectional Signaling Pathway. Arterioscler Thromb Vasc Biol. 2023 Jun;43(6):1015–1030.

